# Genetic Nurture: Estimating the direct genetic effects of pediatric anthropometric traits

**DOI:** 10.1101/2024.12.10.24318796

**Authors:** Samuel Ghatan, Jard de Vries, Jean-Baptiste Pingault, Vincent W Jaddoe, Charlotte Cecil, Janine F Felix, Fernando Rivadeneira, Carolina Medina-Gomez

## Abstract

Parental genetic variants can indirectly influence the traits of their child through the environment, a concept termed "genetic nurture", or indirect genetic effects (IGE). This study estimated the direct genetic effects (DGE), via direct allelic transmission, and IGE shaping height, body mass index (BMI), and bone mineral density (BMD) in a multi-ethnic Dutch pediatric cohort, examining children with repeated measurements at ages six, nine, and thirteen. We imputed missing parental alleles from the phased haplotypes of 1,931,478 variants (MAF >1%), utilizing snipar (single nucleotide imputation of parents). We constructed polygenic risk scores (PRSs) and jointly regressed the proband’s trait on their own PRS, while controlling for the proband’s maternal and paternal PRSs. A total of 4,488 probands, with genetic data, underwent at least one of the three specified measurements. We found statistically significant DGE estimates for the three traits across ages six, nine and thirteen. For instance, 71-77% of the BMI variance explained by the BMI-PRS can be attributed solely to the DGE. IGE estimates reached significance only for BMI measured at ages nine (Beta: 0.05, 95%CI: 0.01-0.09) and thirteen (Beta: 0.05, 95%CI: 0.01-0.09). Maternal and paternal IGE were of a similar magnitude in all our analyses. Our findings indicate that genetic nurture has limited influence on anthropometric traits during formative years. In addition, we do not observe differences between the maternal and paternal indirect contributions to these traits, opposite to the stronger maternal nurturing effect reported for other traits.

## Introduction

Heritability, in the context of genetics, refers to the proportion of phenotypic variance determined by genetic factors [1]. Inherited DNA is passed from parent to offspring during reproduction. Inheritance is governed by Mendel’s laws of genetics, such that each parent passes half of their autosomal DNA to their offspring through meiosis. Consequently, parents and offspring share a resemblance in heritable traits. Parents can also influence the traits of their offspring through the shaping of their offspring’s environment, a phenomenon often referred to as nurture. If the offspring’s trait is influenced by the environment and is heritable, then the genetic variants of the parents can indirectly influence the offspring’s trait via the parent’s own trait and therefore the environment of the child [2]. Thus, the variants of the parent can influence the trait of their offspring through two causal paths: directly by allele transmission and indirectly through the environment. The latter phenomenon was coined “genetic nurture”. Kong et al and Bates et al first demonstrated this concept in a human population by examining the genetic nurture, also known as indirect genetic effects (IGE), for educational attainment [3, 4]. Kong et al showed that the non-transmitted alleles of the parents had an effect on the educational attainment of the proband that is 29.9% (P = 1.6 × 10^−14^) of that of the transmitted polygenic score.We can estimate the direct genetic effects (DGE), i.e. the direct allelic transmission, and IGE by jointly regressing the proband’s phenotype with the polygenic risk score (PRS) of the proband (i.e., child), the mother and the father. A PRS consists of the sum of risk alleles associated with a particular trait, weighted by the effect sizes of those variants as identified in genome-wide association studies (GWAS). In the joint regression model, the DGE are estimated as the magnitude of the effect of the proband’s PRS on the trait of interest, whereas, the IGE are estimated by taking the average effect estimate of the mother’s and father’s PRS within the model [5].

Nurturing conditions play an important role in child development. Growing evidence suggests that early childhood experiences, particularly family environment shape, among others, dietary and physical activity behaviors associated with growth velocity [6, 7]. For example, the adequate intake of basic nutrients through childhood and adolescence plays a crucial role in determining height, accumulation of muscle and fat mass, and accrual of peak bone mass [8–11]. These traits are partially influenced by genetics and are also shaped by the environmental conditions provided by parents. Large GWAS have identified thousands of genetic variants associated with height [12], body mass index (BMI) [13], and bone mineral density (BMD) [14], providing valuable information to estimate an individual’s genetic liability for growth. However, classical GWAS can only estimate the combined effects of the direct and indirect effects [2, 15]. The influence of the family environment on phenotypic variation can change throughout an individual’s life, as interactions among family members evolve. For example, the magnitude of parental nurturing may diminish as a child grows older [16]. It is then plausible that parental IGE, if present, will have the greatest impact on anthropometric traits during childhood and adolescence. To our knowledge, no study has examined the genetic nurturing effect on anthropometric traits in a pediatric population across multiple developmental stages. Therefore, we aimed to estimate the direct and indirect genetic effects in in developmental traits (i.e., height, BMI and BMD) at multiple timepoints through childhood development and test how these effects might vary. Our study surveyed longitudinal data of participants from a Dutch multi-ethnic cohort at ages six, nine and thirteen.

## Methods

### Study characteristics

The Generation R Study is a multi-ethnic population-based prospective cohort study from fetal life onwards based in Rotterdam, The Netherlands. The study recruited pregnant women who were expected to deliver between 2002 and 2006, and encompasses 9,778 mothers and their children [17]. During these years, a vast array of data ranging from detailed questionnaires and regular medical examinations to advanced imaging techniques and extensive biosampling were gathered, and children and parents are in ongoing follow-up. For this study, height, BMI and BMD were measured in the probands during three visits to the research center at a mean age of 6.0, 9.7, and 13.5 years.

### Acquisition of the phenotype data

In the Generation R Study, at each wave of the study, participants removed shoes, heavy clothing, and metal items before measurements. Height was measured to the nearest millimeter by a stadiometer (Holtain Limited, Dyfeld, UK). Weight was measured to the nearest gram using an electronic scale (SECA, Almere, The Netherlands). BMI was calculated [weight (kg)/height (m^2^)]. Height and BMI were then transformed into age- and sex-adjusted standard deviation scores (SDS) using LMSGrowth [18]. Total body BMD was assessed via Dual-energy-X-ray absorptiometry (DXA) scans using a GE-Lunar iDXA device operated by a skilled research assistant with daily quality checks, following standard manufacturer protocols [19]. Total body less head BMD was used for analysis as recommended by the International Society for Clinical Densitometry guidelines [20].

### Genotyping and imputation of the Study population

Probands and parents were genotyped in three batches using three different arrays over a period of 10 years. Due to the limited overlap in overall content across genotyping arrays, the genotype data was first subjected to imputation to increase the number of overlapping variants. Probands were genotyped in two batches. The first round of genotyping was carried out with either the Illumina HumanHap 610 or 660 Quad chips and is referred to here as Generation R3 (GENR3). It comprised data from 5,722 probands after quality control (QC). A detailed description of the genotyping can be found elsewhere [21]. Genotypes were then imputed to the 1000 genomes project phase3v5 (1KGP) reference panel using minimac3. The subset of genome-wide-variants with an r-squared > 0.95 and a minor allele frequency (MAF) > 1%, after imputation, comprised 4,789,340 SNPs.

The second round of proband genotyping, referred to as Generation R4 (GENR4), was conducted together with a subset of participants’ mothers in the study. In total, 3,424 samples (i.e., including both children and mothers) were genotyped in the HuGE facility at Erasmus Medical center. Genome Studio 2.0 was used to initially QC genetic variants and genotype clusters after genotyping. Data was further processed in PLINK 1.9 for the following steps. Exclusion of samples and variants with call rates below 97.5%, variants with an excess of heterozygosity (> 4 standard deviations from the mean), Hardy-Weinberg equilibrium (HWE) deviation (P<1×10^-4^), or samples deemed to be sex mismatches and/or genetic duplicates. In addition, we detected and excluded potential sample swaps by comparing reported familiar relationships based on questionnaires across all Generation R samples (i.e., both children and mothers) and IBD-based relationships. Lastly, zCall was used on the previously uncalled genotypes aiming to improve the call rate of less-frequent to rare variants [22]. More stringent filtering of both samples and variants after zCall filtering was applied including missingness (>99%) and HWE (P<1×10^-7^). In total, 1,809 children and 708,478 variants are included in the final genotyped dataset distributed to Generation R researchers. Next, imputation was carried out. GENR4 variants were surveyed in the 1KG panel, and these not present or with different reported alleles, excluded. The remaining variants were phased using ShapeIT v2 [23], followed by imputation to the 1KG using minimac4 [24] with an in-house pipeline. The subset of variants with an r-squared > 0.95 and MAF > 1%, after imputation, comprised 2,921,550 SNPs.

Parents of probands were genotyped in two rounds, using the GSA-MD v2.0 (n= 1,530 (13%), only mothers), and GSA-MD v3.0 ((n= 10,213 (87%), both mothers and fathers) genotyping arrays. The QC of the first batch is described in detail in the previous section and supplementary figure 1 while the detailed QC of the second batch of GENR Parents is described in figure 2. For this second batch, Genome Studio 2.0 was employed to perform a technical QC of genotyped variants and clusters, followed by a strict PLINK 2 QC. Exclusions were applied to samples and variants with call rates below 97.5%, variants with an excess of heterozygosity (> 4 standard deviations from the mean), HWE deviation (P<1×10^-5^), or samples deemed to be sex mismatches and/or genetic duplicates. In addition, we detected and excluded potential sample swaps by a comparison of reported familiar relationships based on questionnaires across all Generation R samples (i.e., both children and parents) and KING reported relationships [25]. zCall was used on the previously uncalled genotypes aiming to improve the call rate of less-frequent to rare variants [22]. More stringent filtering of both samples and variants after zCall filtering was applied including missingness (>99%) and HWE (P<1×10^-7^). Both sets of parents genotyped data were merged using PLINK 2, and variants not present in both array platforms or reporting different alleles were excluded. In total, the GENR Parents combined information of 660,868 variants present in 11,742 individuals. The combined GENR Parents dataset was phased using ShapeIT v2 [26], followed by imputation to the 1KG reference panel using minimac4 [24] using our in-house pipeline. Imputation resulted in a subset of 3,000,903 SNPs with an r-squared > 0.95 and MAF > 1%. Lastly, retrieved informed consents and family cross-checks based in the four genotyped batches described in the sections above resulted in extra exclusions, noted in an inclusion file share among Generation R researchers. The working genome-wide files for Generation R comprise GENR3 (5,700 participants), GENR4 (1,802 participants) and GENR Parents (11,676, parents).

### Construction of the combined Generation R dataset

The subsets of imputed variant files described above (i.e., MAF > 1%.and r-squared > 0.95), that were present across all three files, were then merged. Palindromic variants (coded as A-T or G-C) with ambiguous effect allele frequencies (40-60%) were removed. Variants that exhibited batch effects, as determined by examining PCA plots and by conducting a GWAS using batch as the outcome, were subsequently removed. Overall, the merged set comprised 1,931,478 variants and 19,178 samples after QC. This merged set of variants was then phased with SHAPEIT2 [26] with the duohmm function, -W 5 parameter (window size) and the 1KGP reference panel (All population) as a genetic map [27]. The duohmm function utilizes pedigree information to improve phasing.

### Determination of the Genetic ancestry of the participants

Individuals of European genetic ancestry were identified using genetic data from the 1KGP as a reference. Our dataset and the European set of the 1KGP (e.g., CEU, TSI, GBR, FIN, IBS) were harmonized to ensure overlapping variants and consistent allele coding. Palindromic variants and variants with MAF < 1% were removed. In the merged dataset, variants were then pruned (r2>0.2 in a 50kb window). Principal component analysis (PCA) was conducted using PLINK. Using the first two principal components, the center of the 1KGP European samples was calculated. The maximum Euclidean distance of the 1KGP European samples was then calculated. All GENR samples whose maximum Euclidean distance fell beyond this radius were defined as non-European.

### Imputation of missing parental alleles

DGE and IGE can only be estimated in complete trios. In duos, the missing/ungenotyped parental alleles have to be imputed. We utilized snipar (single nucleotide imputation of parents) to impute missing parental alleles in probands of European ancestry [5]. Briefly, snipar imputes ungenotyped parental alleles based on Mendelian laws of inheritance. Feasible nonlinear imputations of parental alleles can be grouped into three categories: (a) genotyped sibling pairs, (b) genotyped parent–offspring pairs, and (c) genotyped sibling pairs with one genotyped parent. By using phased genotype data, uncertainty at heterozygous positions can be resolved. Missing genotypes that cannot be inferred through family structure were imputed using the in-sample population allele frequencies. As a result, only parents of European ancestry were included in the imputation. Phased VCF files were converted to bgen files using QCtools. Identity by decent (IBD) segments shared between siblings were inferred using the implementation within snipar, which employs a Hidden Markov Model [5]. KING software [25] was used to obtain kinship coefficients and pedigree information. A kinship coefficient of 0.177 and above was used to define first degree relatives. KING identified 10,229 parent offspring relations. Families consisted of 3,635 trios and 2,897 duos and 267 proband-sibling pairs. Kinship files, as well as files denoting the age and sex of individuals were used to infer IBD segments and pedigree structure. Missing parental genotypes (Table were imputed by snipar for 1,931,478 variants using phased haplotypes. After imputation and QC a total of 5,241 European and non-European probands had complete parental data available (Table 1).

**Table 1.** Overview of the familial data types within the Generation R Study.

| Data type | N probands | Missing data imputed |
| --- | --- | --- |
| Proband and both parents (European) | 2,736 | No |
| Proband and one parent (European) | 1,576 | Yes |
| Proband and sibling(s) but no parents (European) | 57 | Yes |
| Proband and both parents (Non-European) | 872 | No |
| Total | 5,241 |  |

### Polygenic risk scores

To construct the PRS for each trait, variant effect estimates were obtained from GWAS summary statistic data for height [12], BMI [13], and estimated bone mineral density (eBMD) [14]. These GWAS were chosen due to their representation of the largest sample sizes available for each respective trait. To increase the variance explained by each PRS, LDpred2 [28] was used to construct genome-wide PRSs across autosomal chromosomes. LDpred2 uses a Bayesian approach that adjusts the effect estimates of GWAS summary statistics using linkage disequilibrium (LD) information to estimate a posterior mean effect size for each SNP, resulting in a more accurate PRS prediction. The LDpred automatic model was used, in which hyper-parameters are directly inferred from the data without the need for a validation set. Variants in the GWAS summary statistics were intersected with the 1,931,478 imputed variants of our sample and the 1,444,196 “HapMap3+” variants [29]. Palindromic variants were removed. In sample LD matrices were generated from the imputed variants of the probands. Once variant weights had been computed using LDpred, snipar was used to generate the PRSs for the probands, mothers and fathers. The same set of variants was used to construct the proband and parental PRSs. To obtain estimates of direct and indirect genetic effects, we jointly regressed the proband’s trait on their own PRS, while controlling for the proband’s maternal and paternal PRS. IGE are quantified as the average of the maternal and paternal PRS effects within equation 1.

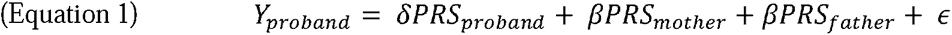

Where, *Y* = proband phenotype, δ = direct genetic effect, *β*(*PRS_mother_* + *PRS_father_*)/2 = indirect genetic effect. Linear regression models were constructed using R software and were adjusted by age, age^2^, sex, the interaction between age and sex, and principal components 1 to 10. Principal components were constructed from the genetic data of the individuals being tested. Traits were standardized to have a mean of zero and a standard deviation of one. The PRS for eBMD and BMI was tested in the European subset of the sample, as the GWAS for these traits was derived from a European population and was poorly associated within our non-European sample. The height GWAS from which summary statistics were extracted was performed in a multiethnic population and as such, we used the complete GENR dataset to infer DGE and IGE. A two-sided Z-test was used to calculate the difference in effects between the maternal and paternal PRSs, as well as, the difference in effects between different age measurements. The ratio of IGE and DGE can be determined using the following method: DGE/IGE+DGE. To determine the fraction of the phenotype variance which is explained by the DGE we square this ratio.

### Analysis in independent probands

Closely related individuals can cause inflation in the estimates of the association of the PRS and the trait tested [30]. This phenomenon arises, in part, because individuals from the same family share genetic information but also more environmental factors than unrelated individuals. For instance, individuals in nuclear families often share the same household. If these shared factors are associated with the trait of interest, it can lead to confounding in PRS analyses [30]. While GWAS methods attempt to control for broad population stratification, they may not adequately control for subtle stratification caused by family relationships [31]. To evaluate whether the presence of related participants of the Generation R Study led to inflated association effect sizes, we performed a sensitivity analysis in which children related up to the third degree (i.e., first cousins) were removed from the regression analysis. Kingship coefficients were derived from the analysis performed in KING.

### Re-imputation of observed parental genotypes

To assess the quality of the imputation we proceeded to re-impute measured parental alleles and then compared the re-imputed alleles to the observed alleles. To perform the analysis, we started with complete trios families of European ancestry (n = 8055), of which 2,987 were probands. 5,470 were parents. Next, the allele information of one parent from 844 probands (30%) was randomly removed. Missing parents genotypes were imputed using snipar and then the correlations between imputed genotypes and measured genotypes were tested for each individual across 1,931,478 variants. The mean correlation across all individuals and variants was as our measure of the imputation accuracy.

## Results

### Description of the data acquisition

Proband’s measurements were collected during three visits to the research center, roughly four years apart. Measurements of probands took place at the median age of 6.0 (IQR: 5.9-6.2), 9.73 (IQR: 9.6-9.9), and 13.5 (IQR: 13.4-13.7). Of the 4,177 probands that had one of the three measurements and genotype data available, 2,587 (62%) had measurements across all three visits. Between 83-84% of the participants measured at each visit, were of European ancestry. A full description of the participants taking part in each measurement wave can be found in Table 1.

### Imputation of missing parental genotypes

In total 1,690 missing parental genotypes of European ancestry were imputed using snipar (Table 2). The structure of the missing family data consisted of 1,319 missing father genotypes with the mother and proband genotypes available, 257 missing mother genotypes with the father and proband genotypes available, and 57 probands with no parental genotype data but with a sibling genotyped. To assess the quality of the imputation we re-imputed measured parental alleles and calculated the correlation between the re-imputed alleles and observed alleles. When assessing the quality of the imputation, the mean correlation between the observed alleles and the re-imputed alleles was 0.82, indicating a small bias in the imputed data.

**Table 2.** Descriptive statistics of the individuals contributing data for this study at ages, six, nine and thirteen.

| Characteristic | Measurement age 6 | Measurement age 9 | Measurement age 13 |
| --- | --- | --- | --- |
| Probands with family genetic data | 5241 | 5241 | 5241 |
| Attended assessment center | 4178 | 3831 | 3232 |
| Age (years) | 6.01 (5.86-6.23) | 9.73 (9.61-9.87) | 13.54 (13.42-13.71) |
| Female (%) | 49.42 | 49.64 | 49.22 |
| European (%) | 84.38 | 84.4 | 83.2 |
| BMI (sds) | 0.17 (-0.34-0.75) | 0.18 (-0.48-0.87) | 0.14 (-0.56-0.90) |
| NAs | 1 | 74 | 6 |
| Height (sds) | -0.18 (-0.84-0.47) | -0.09 (-0.70-0.57) | 0.06 (-0.63-0.74) |
| NAs | 1 | 74 | 6 |
| TBLH-BMD (g/cm<sup>2</sup>) | 0.55 (0.52-0.58) | 0.67 (0.64-0.72) | 0.85 (0.78-0.92) |
| NAs | 108 | 117 | 176 |

### Estimating direct and indirect genetic effects for childhood anthropometric traits

The PRS for BMI was constructed using 195,154 genetic variants across the genome and explained between 7.7-14.0% of the BMI phenotypic variance (supplementary table 1) at each measurement (European subset). We observed a correlation coefficient (ρ) of 0.55 (95%CI: 0.53-0.57) between proband and mother PRS, a ρ of 0.57 (95%CI: 0.55-0.59) between proband and father PRS, and a ρ of 0.06 (95%CI: 0.02-0.10) between the parent’s PRSs. We obtained statistically significant DGE estimates for BMI across all ages (Figure 1a, supplementary table 1). DGE explained between 6.0-10.5% of the BMI phenotypic variance (supplementary table 1) at the different ages the assessment was performed. Conversely, IGE estimates on BMI reached statistical significance for the measurements at age 9 (Beta: 0.05, 95%CI: 0.01-0.09) and at age 13 (Beta: 0.05, 95%CI: 0.01-0.09). The IGE estimate at age 6 was borderline significant (Beta: 0.03, 95%CI: -0.01-0.07). The ratio of DGE/IGE+DGE ranged between 0.84-0.88 across the different ages. Between 71-77% of the total variance explained by the BMI PRS is due to the direct effects alone. There were no significant differences between maternal and paternal effects or between age groups at any of the ages tested.

**Figure 1.**
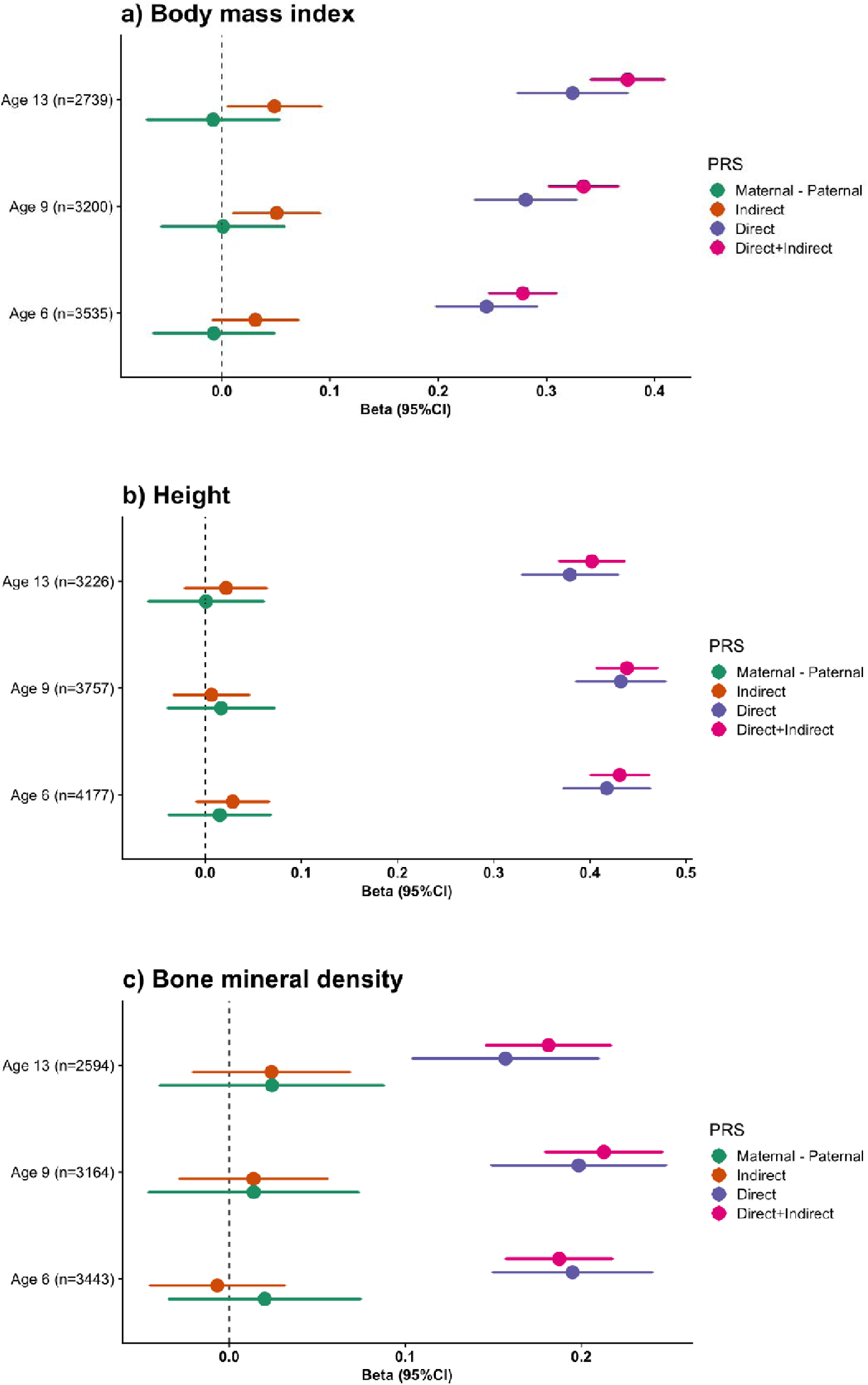
Forest plots depicting the direct, indirect and maternal minus paternal genetic effects for: a) Body mass index b) Height c) and Bone mineral density measured at different ages in participants of the Generation R Study.

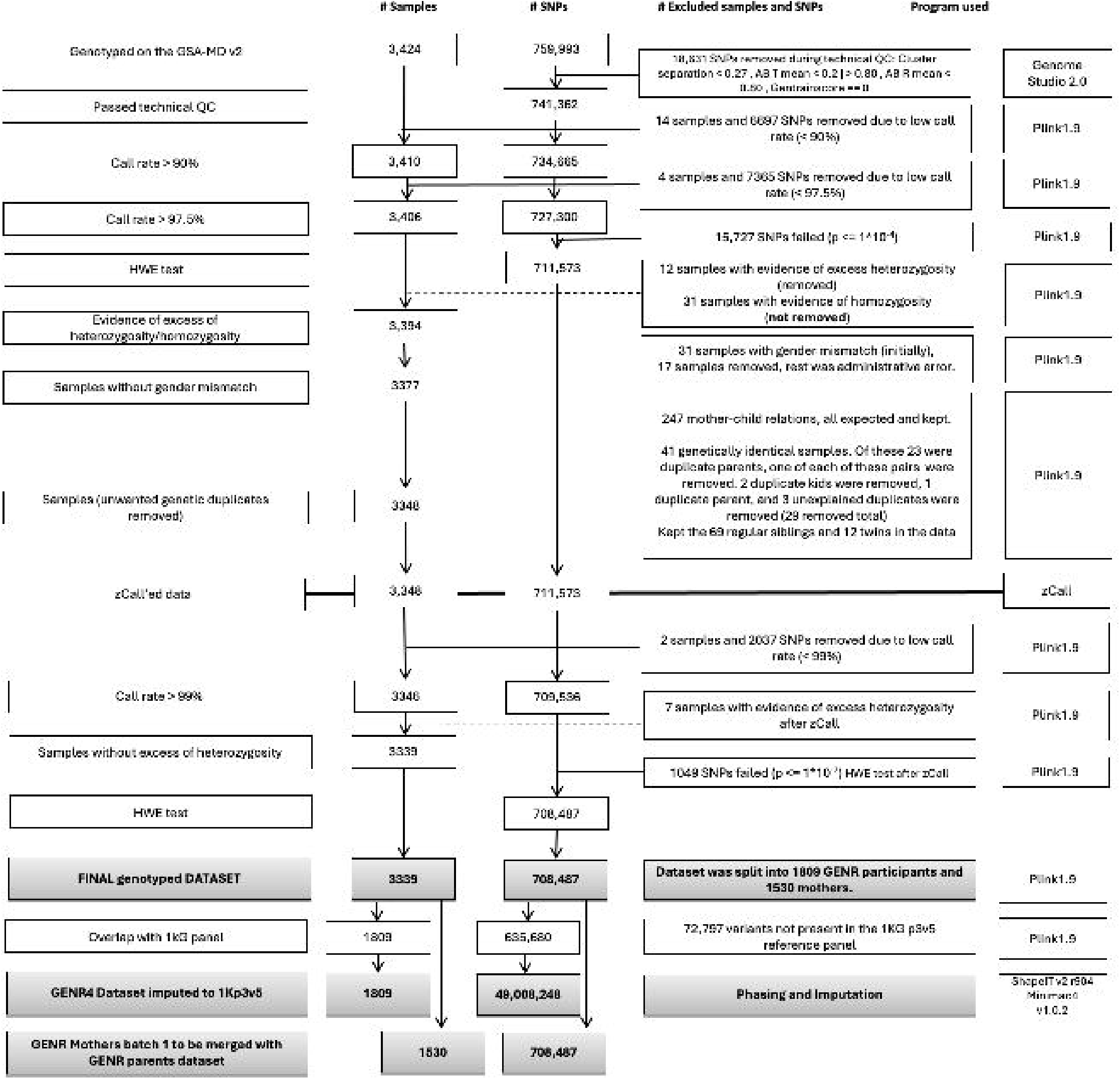
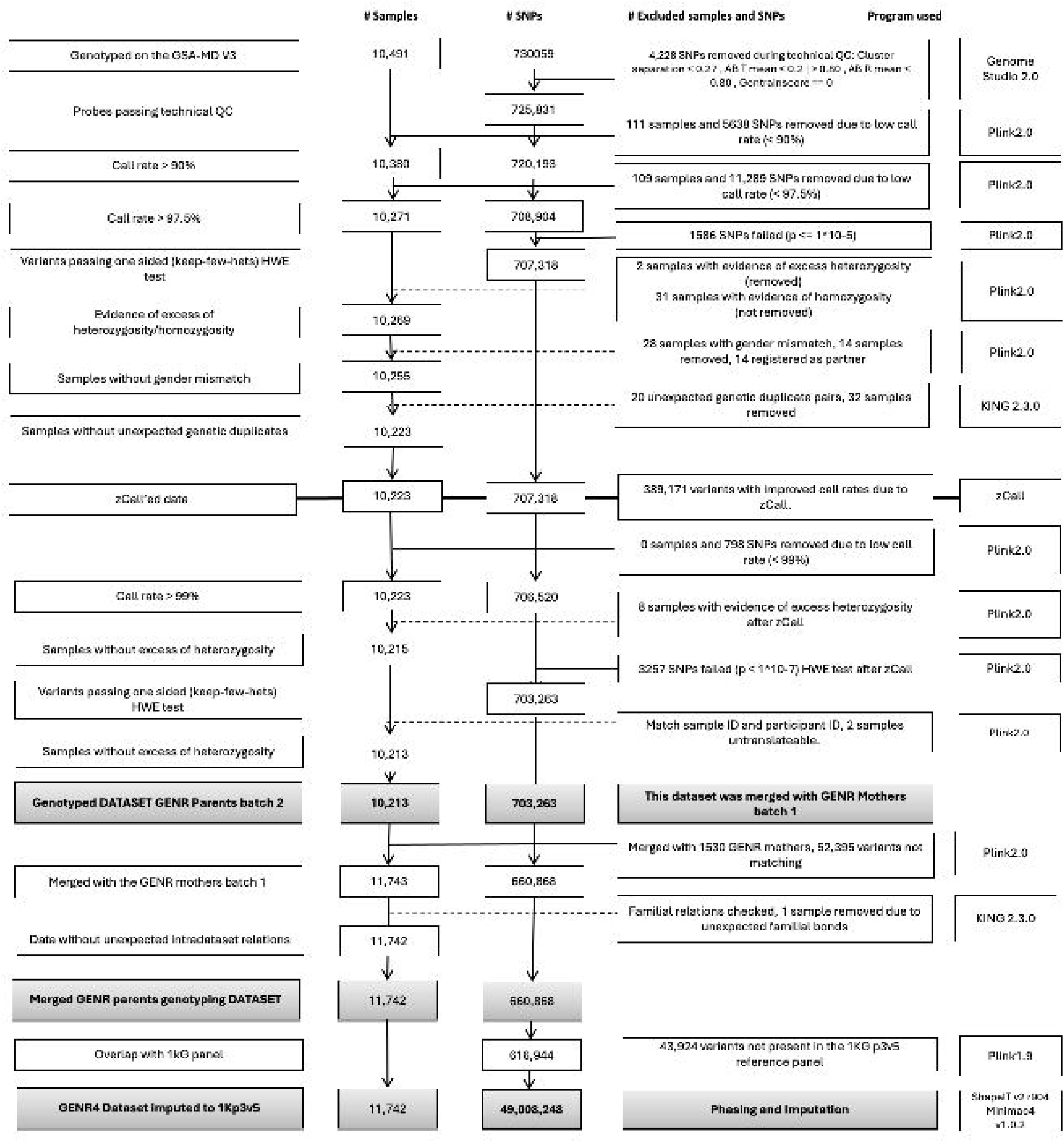

The height PRS was constructed using 210,390 genetic variants across the genome and explained between 16.2-19.2% of the phenotypic variance of height (supplementary table 1) at each measurement wave. The correlation between proband and mother PRSs was 0.60 (95%CI: 0.59-0.61), proband and father PRSs, 0.62 (95%CI: 0.61-0.63) and between parent’s PRSs, 0.17 (95%CI: 0.16-0.18). We obtained statistically significant unbiased DGE estimates for height across all ages (Figure 1b, supplementary table 1). DGE explained between 14.4-18.7% of height phenotypic variance (supplementary table 1). IGE Estimates of did not reach significance for any of the ages tested (p-value > 0.05). Direct effects explained between 94-99% of the height variance explained by its correspondent PRS. There were no significant differences between maternal and paternal effects or between age groups at any of the ages tested.

The PRS for BMD was constructed using 251,272 variants across the genome and explained between 3.3-4.5% of the total body less head BMD phenotypic variance (supplementary table 1) at the different measurement waves (European subset). The correlation between proband and mother PRSs was 0.53 (95%CI: 0.51-0.55), proband and father PRSs were 0.56 (95%CI: 0.54-0.58) and between parent’s PRSs was 0.04 (95%CI: -0.01;0.06). We obtained statistically significant unbiased DGE estimates for BMD across all age groups (Figure 1c, supplementary table 1). DGE explained between 2.5-3.9% of the BMD phenotypic variance (supplementary table 1). IGE estimates did not reach statistical significance for any of the ages tested. There were no significant differences between maternal and paternal effects or between age groups at any of the ages tested.

### Sensitivity analysis

To assess the extent to which our PRS coefficients were inflated due to relatedness within our sample population, we conducted a sensitivity analysis including only independent individuals (supplementary table 2). Overall, DGE and IGE coefficients remained consistent between both analyses indicating minimal bias due to relatedness in our analysis (supplementary table 2).

## Discussion

We examined genetic nurture effects that may shape childhood anthropometric traits across ages six, nine and thirteen in a multi-ethnic Dutch cohort study. We imputed missing parental genotypes based on Mendelian inheritance. Polygenic risk scores (PRSs) were constructed for mothers, fathers and probands, for three childhood anthropometric traits: height, BMI and BMD (total body less head), using hundreds of thousands of genetic variants. We obtained statistically significant estimates of direct genetic effects (DGE) for all traits. We also observed significant estimates of indirect genetic effects (IGE) for BMI but not height or BMD. In general, we observed modest estimates of genetic nurture effects of anthropometric traits throughout developmental ages, with no significant differences in IGE for any traits between different age ranges in children. Additionally, no differences were observed in the contribution of maternal and paternal genetic nurture effects. These results indicate that the genetic effects of anthropometric traits in children predominantly occur via genetic transmission, although some genetic nurture effects are present for BMI.

Our study is not the first to examine the effects of genetic nurture on child BMI trajectories. Tubbs et al. examined the influence of maternal genetic nurture on child BMI trajectories in approximately 2,900 children and found that its effect increased with age, becoming stronger throughout development [32]. Notably, Tubbs et al. focused solely on maternal genetic nurture, without accounting for potential paternal influences. This finding contrasts with studies on twins, which suggest that the impact of the shared environment, including genetic nurture, diminishes over childhood and adolescence [33]. Further complicating the picture, a GWAS of BMI at different ages found that distinct genetic factors influence BMI during infancy (2 weeks to 18 months) compared to later childhood (18 months to 13 years) [34]. Similarly, a study based on pooled twin data reported that genetic correlations for BMI decrease with age, indicating that new genetic factors emerge at different stages of childhood growth [35]. In the Generation R study, a polygenic risk score (PRS) for adult BMI explained varying amounts of BMI variance across age groups, with the highest variance at age 13, suggesting that different genetic variants influence BMI across childhood, or that adult BMI variants have weaker effects at younger ages. Additionally, a study tracking BMI PRSs from childhood to adulthood found that differences in weight between individuals in the highest and lowest deciles of the PRS were evident early in childhood and grew to 12 kilograms by age 18 [36]. Together, these findings highlight the dynamic role of genetic factors, with varying influences across different developmental stages.

In a prior study that assessed genetic nurturing effects in 21,637 adult probands from an Icelandic cohort study, a DGE/IGE+DGE ratio of 0.94 for height was reported [3] aligning with the estimated ratios from our study. Despite the large sample size of the Icelandic study, the authors did not find a significant IGE for BMI [3]. In contrast, we report a significant IGE for BMI. As the Icelandic study was carried out on adults, it is plausible that the influence of the IGE on BMI is more accentuated during childhood. This aligns with the logic that parents have more control over their child’s environment during childhood, but less control over their environment in adulthood. Still, we observed no significant disparities in IGE on BMI across the developmental ages of our examined cohort, suggesting that a potential divergence of IGE may occur later in adolescence or adulthood. Another study of roughly 56,500 adult probands from the UK biobank, Generation Scotland and the Swedish Twin registry produced a DGE/IGE+DGE ratio of 0.910 (s.e. = 0.009) for height, and 0.962 (s.e. = 0.017) for BMI [37]. Again, the IGE values for height in this study are similar to those obtained from the Generation R Study, while the IGE values for BMI in this study are lower. However, the Generation R Study displays larger standard errors for BMI IGE and the BMI GWAS summary statistics used in our study were different. No prior studies have investigated the IGE or DGE of BMD in either pediatric or adult populations, precluding a direct comparison with our research. While IGE for BMD did not reach statistical significance, we did observe increasing effect estimates with age. Correlations between parental scores for height, and to a lesser extend BMI, were inflated, indicating bias, possibly driven by assortative mating or population stratification, which are also captured by IGE [39, 40]. Assortative mating refers to the phenomenon in which individuals are more likely to mate with those who share a similar trait than is expected under random mating, and is a well-documented phenomenon of the genetics of height [5, 39, 40]. Previous studies have shown that much of the IGE on height is attributable to assortative mating, whereas, for BMI, assortative mating has been shown to have a negligible effect [37, 40]. BMD on the other hand displayed no evidence of assortative mating in our study, in accordance with previous estimations [39]. We did not observe any statistically significant difference between maternal and paternal IGE on the outcome traits across the age groups tested. We are underpowered to draw reliable conclusions regarding parental differences in genetic nurturing effects. Yet, our results do not support the hypothesis that mothers have a stronger nurturing effect on children’s anthropometric traits during their formative years, or the hypothesis that IGE for anthropometric phenotypes are mainly driven by interactions between the mother and fetus in the womb for the PRS that we studied. However, the nurturing effects of the mother may still influence the child via alternative environmental pathways.

The use of PRS derived from GWAS, which are inherently influenced by indirect genetic effects, can have clinical significance, especially when using PRS for disease screening [41, 42]. Screening children using a PRS comprised of DGE would offer a more accurate classification of children suffering from a possible Mendelian disease, or accumulation of common risk alleles, than using a PRS consisting of both DGE and IGE [43]. Further, one potentially promising use of PRS is for personalized medicine, in which individuals with a high genetic risk of obesity would be prescribed medication for earlier prevention and treatment, whereas individuals with a low genetic risk of obesity would be prescribed lifestyle interventions [36]. In such a scenario, it is of importance to construct a PRS without IGE to distinguish those with a high genetic burden of disease from those with a high environmental burden. Additionally, IGE can lead to biases within Mendelian randomization, a type of causal association analysis, leading to spurious associations [44]. IGE can also have impacts on many downstream analyses of GWAS, including annotation, heritability estimates and genetic correlations [5].

Our study presents several limitations. The limited sample size of our cohort warrants replication of our results in additional (larger) cohorts to improve power and generalizability of the effect estimates. Although our study encompassed multiple ethnicities, we could not conduct multi-ethnic analyses for BMI and BMD due to the absence of sufficiently large multi-ethnic GWAS summary statistics available. Additionally, current BMD GWAS have a limited sample size affecting power, therefore we constructed a BMD PRS derived from an eBMD GWAS. Despite also being a measure of bone mineralization, this measurement is only moderately correlated with the BMD obtained from dual-energy x-ray absorptiometry, which was measured in our cohort [14]. Lastly, our estimates of IGE may be inflated due to assortative mating, particularly for height. Here, we estimated DGE and IGE using PRSs derived from GWAS of unrelated individuals [12–14]. Therefore, the IGE could reflect biases, such as population stratification and assortative mating, present within the original GWAS. An improved approach would be to use variant effect estimates derived from family-based GWAS design, in which parental genotypes are controlled for when testing the association between proband genotypes and traits, which do not suffer from these biases [2, 15, 45]. However, such family-based GWAS designs would require large samples of genotyped nuclear families, currently not available.

In conclusion, we observed moderate estimates of genetic nurturing effects on height, BMI and BMD throughout developmental ages, with no significant differences in indirect genetic effects for any of the traits across different ages in children. Additionally, no differences were observed in the contribution of maternal and paternal genetic nurturing effects, providing no indication of non-genetic transmission of the mother’s anthropometric traits on the child’s in pregnancy. These results do not support the hypothesis that mothers have a stronger nurturing effect on children during their formative years, or the hypothesis that IGE for anthropometric phenotypes are mainly driven by interactions between the mother and fetus in the womb.

## Data Availability

All data generated or analyzed during this study are included in this published article and its supplementary information files.

## List of abbreviations

(GWAS): Genome-wide association study
(LD): linkage disequilibrium
(PRS): polygenic risk score
(BMI): body mass index
(BMD): Bone mineral density
(DGE): Direct genetic effects
(IGE): Indirect genetic effects.

## Declarations

Not applicable.

## Ethics approval and consent to participate

The Generation R Study was approved by their respective institutional ethics review committees and all participants provided written informed consent.

## Consent for publication

Not applicable.

## Data and code availability

Height and BMI summary statistics can be obtained from GIANT consortium website (https://portals.broadinstitute.org/collaboration/giant/index.php/GIANT_consortium_data_fil es). Estimated bone mineral density summary statistic data can be obtained via the GEFOS consortium website (http://www.gefos.org/). All data generated or analyzed during this study are included in this published article and its supplementary information files.

## Authors’ contributions

S.Ghatan, C.Medina-Gomez, and F.Rivadeneira designed the study. S.Ghatan performed the analysis. S.Ghatan, F.Rivadeneira, and C.Medina-Gomez drafted the manuscript. All authors contributed to the interpretation of data and critical revision of the manuscript. All authors read and approved the final version of the manuscript.

## Acknowledgments

We express our gratitude to the study participants, whose participation made this work possible, and to the numerous colleagues who contributed to the collection, phenotypic characterization of clinical samples, as well as genotyping and analysis of GWAS data. We gratefully acknowledge the contribution of children and parents, general practitioners, hospitals, midwives and pharmacies in Rotterdam. The Generation R Study is conducted by Erasmus MC, University Medical Center Rotterdam in close collaboration with the School of Law and Faculty of Social Sciences of the Erasmus University Rotterdam, the Municipal Health Service Rotterdam area, Rotterdam, the Rotterdam Homecare Foundation, Rotterdam and the Stichting Trombosedienst & Artsenlaboratorium Rijnmond (STAR-MDC), Rotterdam. The study protocol was approved by the Medical Ethical Committee of the Erasmus Medical Centre, Rotterdam. Written informed consent was obtained for all participants. The generation and management of GWAS genotype data for the Generation R Study was done at the Human Genomics Facility, HuGe-F, housed within the Laboratory for Population Genomics of the Department of Internal Medicine at Erasmus MC. Genetic Laboratory of the Department of Internal Medicine, Erasmus MC, The Netherlands. We thank Pascal Arp, Karol Estrada, Mila Jhamai, Manoushka Ganesh, Gaby van Dijk, Marijn Verkerk, Lizbeth Herrera, Marjolein Peters, and Dr. Linda Broer for their help in creating, managing and QC the GWAS database.

